# Factors linked to changes in mental health outcomes among Brazilians in quarantine due to COVID-19

**DOI:** 10.1101/2020.05.12.20099374

**Authors:** Matthew Stults-Kolehmainen, Alberto Filgueiras, Miguel Blacutt

## Abstract

This aim of this investigation was to track changes and risk factors for mental health outcomes during state-mandated quarantine in Brazil. Adults residing in Brazil (n = 360, 37.9 years old, 68.9% female) were surveyed at the start of quarantine and approximately three weeks later. Outcomes assessed included perceived stress, state anxiety and symptoms of depression. Aside from demographics, behaviours and attitudes assessed included exercise, diet, use of tele-psychotherapy and number of COVID-19 related risk factors, such as perceived risk of COVID-19, information overload, and feeling imprisoned. Overall, all mental health outcomes worsened from Time 1 to time 2, although there was a significant gender x time interaction for stress. 9.7% of the sample reported stress above the clinical cut-off (2 SD above mean), while 8.0% and 9.4% were above this cut-off for depression and anxiety, respectively. In repeated measures analysis, female gender, worsening diet and an excess of COVID-19 information was related to all mental health outcomes. Positive dietary changes were associated with decreases in depression and anxiety. Exercise frequency was positively related to state anxiety and perceived stress (0 days/week > 6 days/week). Those who did aerobic exercise did not have significantly increase in depression. Use of tele-psychotherapy predicted lower levels of depression and anxiety. In multiple regression, anxiety was predicted by the greatest number of COVID-19 specific factors. In conclusion, mental health outcomes worsened for Brazilians within the first month of quarantine and these changes are associated with a variety of risk factors.

## Introduction

Mental health consists of the set of emotions, thoughts and behaviours that enable individuals to work, cope and deal with problems in everyday tasks (World Health Organization, 2004). Historically, although researchers from the biomedical sciences dedicated more time and resources in the study of physical health, findings from the last 50 years have slowly captured the interest of scientists from diverse fields to look upon mental health to explain somatic diseases, physical functioning, quality-of-life, well-being and work productivity, (Christensen et al., 1999; Prince et al., 2007; Stults-Kolehmainen et al., 2014). For instance, mental health is associated with disability-adjusted life years (DALYs) and premature mortality (Vigo et al., 2019) with 17% of DALYs attributable to mental health in Brazil and 22% in the United States. Those with worse mental health, such as higher levels of chronic stress, have a greater risk for physical health problems, such as cardiovascular disease (Stults-Kolehmainen, 2013). Poor mental health costs society a significant amount of money, in terms of lost productivity, strain on healthcare systems, loss of income and other consequences (Trautmann et al., 2016). Conversely, recent research from the World Health Organization suggests that every American-dollar spent in mental health care is equivalent to a return of four American-dollars in better well-being and ability to work (Wilson, 2016). Thus, a person with good mental health is likely physically healthy, happy, productive, and can contribute to a greater functioning of society (Prince et al., 2007; Wilson, 2016).

The recent outbreak of the Coronavirus Disease 2019 (COVID-19), caused by SARS-CoV-2, around the world at the end of 2019 and the beginning of 2020 led to a series of guidelines to avoid mass contamination and limit its lethality (World Health Organization, 2020). Quarantine, confinement, and social-distancing have been common measures taken to limit the spread of COVID-19 (Wilder-Smith and Freedman, 2020). As a result, for periods of time people have not been able to freely from their homes, they have needed to keep a 2-meter physical distance from one another on the streets, and infected people are obliged to be confined in hospitals or their own homes without any kind of physical proximity to others. These restrictions are intended to benefit the physical health and safety of all people and it is widely accepted that these regulations save lives. However, these measures have come at a cost to the mental health and well-being a substantial proportion of the population (Rubin and Wessely, 2020). Furthermore, not all individuals in Brazil adhered to quarantine guidelines when they were in force Obedience of Brazilians to social isolation during quarantine peaked at 63% on March 23^rd^ 2020 and dropped to 47% in April 2020 (INLOCO, 2021) perhaps explaining why Brazil had the highest contagion rate (R0 = 2.81) in the world during April, 2020 (Imperial College COVID-19 Response Team, 2021).

An updated systematic review on the effects of social distancing and quarantine on mental health revealed that anxiety, depression, stress, anger, insomnia, hopelessness, and sadness were all increased during those conditions (Brooks et al., 2020). A recent study (Hu et al., 2020) from a cross-national sample (n = 992) in China found that state-anxiety scores significantly increased as strictness of quarantine increased across the following conditions: no quarantine, closed community, voluntary quarantine, fully closed community, forced quarantine at home, medical observation, and centralized quarantine. Other behavioural problems also appear during this period; participants in a nationwide survey recently published in China reported nutritional issues, lack of ability to exercise and numerous changes in daily routines and habits (Qiu et al., 2020). Accordingly, psychosocial and behavioural dimensions seem associated under quarantine conditions (Filgueiras and Stults-Kolehmainen, 2021; Blacutt et al., 2021). Similar findings were also depicted in research conducted in other quarantine situations, such as: the Severe Acute Respiratory Syndrome (SARS) epidemic in Canada (Hawryluck et al., 2004), Taiwan (Bai et al., 2004) and Hong Kong (Lee et al., 2005), the Middle East Respiratory Syndrome (MERS) epidemic caused by another strain of coronavirus in Korea (Jeong et al., 2016) and the equine influenza epidemic in Australia (Taylor et al., 2008). Altogether, the evidence suggests that quarantine leads to an increase of mental health problems. Specific to Brazil, Blacutt et al. (2021) recently found significant increases in the incidence, prevalence, and severity of stress, depression, and anxiety in the early phases of the COVID-19 pandemic in a sample of people from Brazil.

Identifying risk factors that modify the mental health experience of quarantine and social isolation is important. Research among people in normal and healthy conditions has shown that sociodemographic variables, health behaviours and other daily routines are linked to better mental health. Among the most commonly investigated demographic variables are gender (Almeida and Kessler, 1998; Nolen-Hoeksema and Hilt, 2009), education (Steele et al., 2007) and age (Christensen et al., 1999). For health behaviours, a large literature suggests that moderate to vigorous physical exercise from three to five times per week leads to reduced anxiety (Wipfli et al., 2008), depression (Craft and Landers, 1998; de Oliveira et al., 2018), stress (Stults-Kolehmainen and Sinha, 2014) and other mental health issues (Landers and Arent, 2007). Similar associations are found with dietary habits; a diet low in fat, sugar or carbohydrate tends to be associated with fewer psychological issues (Molendijk et al., 2018; O’Neil et al., 2014). Aside from these health behaviors, finding and receiving mental health support is imperative for many individuals at risk. Psychologists and other mental health practitioners who provide online or tele-psychotherapy may also help to improve mental health conditions (Varker et al., 2019).

Unfortunately, resources are scarce in every field of the health system, including those for mental health (Qiu et al., 2020). Therefore, it is pivotal to establish a priori where and how to invest those scarce resources. This is a difficult task because the current stressor is highly unique. Quarantine is due to a pandemic of truly global proportions that has reached every level of society, with a long but discontinuous and intermittent duration, resulting in remarkable social upheaval (World Health Organization, 2020). There is little research on the association between psychological, demographic and behaviour variables in the general population during society-wide social isolation. Furthermore, it is a consensus that psychological phenomena, such as stress and depression, are multifactorial in their etiology and manifestation, with a large amount of variables to consider (Wilson, 2016; World Health Organization, 2004). In order to help governments, service providers and scientists to establish public policies toward resource allocation in mental health during the continued COVID-19 pandemic crisis, this study aimed to fill the gap in the current literature. Three psychological dimensions were queried due to their relevance in the literature: (i) perceived stress (Hawryluck et al., 2004; Qiu et al., 2020), (ii) depression (Brooks et al., 2020) and (iii) state anxiety (Jeong et al., 2016; Rubin and Wessely, 2020). The aims of this investigation were two-fold. First, this research was intended to track mental health changes over two time points during quarantine. The second objective was to associate mental health outcomes with pertinent demographic, behavioural and COVID-19 specific factors.

## Materials & Methods

The present research is a longitudinal psychosocial study that collected data in two periods: the first week of state-mandated quarantine (São Paulo, 2020) and approximately three weeks after this mandate. The Ethical Committee of the first author’s institution approved the project under the process #2020.2014-0932-12. Participants were allowed to leave the online questionnaires at any time and procedures obeyed the Declaration of Helsinki.

Volunteers were recruited through a snowball sampling method, which started by posting messages about the research study on social media (Heckathorn, 2011). We recruited 360 (248 women, 68.9%) Brazilians or foreigners living in Brazil from 9 States and 23 different cities. sThis research was conducted in Brazilian Portuguese, so it was necessary to know how to read and write in this language. All participants digitally signed the Term of Consent and agreed to be contacted after the first round of data collection to be part of the second round.

There were four instrument measures adopted: a sociodemographic and attitudinal questionnaire. Mental health outcome variables were assessed at Time 1 and Time 2, these included the Perceived Stress Scale with 10 items (PSS-10), the Filgueiras Depression Inventory (FDI) and the State subscale of the State-Trait Anxiety Inventory (S-STAI). At Time 1, participants additionally answered a 9-item sociodemographic questionnaire, which included the following question in this order: (i) age, (ii) gender, (iii) education, (iv) height, (v) weight, (vi) whether the participant had any physical risk factor for COVID-19, and whether he/she used one of the following during quarantine: (vii) telepsychotherapy, (viii) telemedicine, (ix) online nutritionist and/or an online fitness coach. At Time 2, participants were asked about their quarantine nutrition, exercise habits, and quarantine information, which included the following questions: (i) frequency of exercise during quarantine in days, (ii) whether there were changes in the frequency of exercise comparing before and during quarantine (“no changes”; “increased exercise frequency” and “decreased exercise frequency”) and (iii) types of exercise (“aerobic”; “anaerobic”; “both”; or “no exercise”), (iv) change in diet (“for the worse”; “neutral”; or “for the better”), (v) weight gain during quarantine (“none”; “less than 5kg”; or “more than or equal to 5kg”), and (vi) weight loss (“none”; “less than 5kg”; or “more than or equal to 5kg”),). Additionally at Time 2, participants were asked about: (vii) the amount of COVID-19 information the participant felt he/she was receiving (“Too much information”; “Enough information”; or “Little information”). Another three items were provided on a five-point Likert-type scale ranging from 1 “Totally agree” to 5 “Totally disagree”; the items were: (viii) “Do you feel imprisoned due to this quarantine?”, (ix) “Do you feel you are able to understand what is happening?”, (x) “Do you trust your own ability to differentiate good from bad sources of information?”.

The PSS-10 (Cohen and Williamson, 1998) is a 10-item questionnaire that asks individuals about their perceptions regarding stress-like symptoms. It is answered on a five-point Likert-type scale ranging from 0 “Never” to 4 “Very often” (scores range from 0-40). The population mean is 17.0 (SD = 5.02) with a score over 27 indicating excessive stress (Cacciari et al., 2016). The FDI (Filgueiras et al., 2014) is a 20-item scale that asks individuals to grade the level of association between the respondent’s own self-perception and one-word items extracted from depression symptoms listed in the DSM-V in the last fortnight. It is rated on a six-point Likert-type scale ranging from 0 “not related to me at all” to 5 “totally related to me” (scores range from 0-100). The reference mean is 53.3 (SD = 17.3) with 88 or higher indicating a cut-off for depressive symptomology (Filgueiras et al., 2014). The S-STAI (Spielberger et al., 1983) is a subscale of a broader questionnaire that assess state (i.e., one’s current mood state) and trait (i.e., dispositional and personality-related traits) anxiety. The focus of S-STAI is the mood state of the respondents who answer questions about her/his own feelings on a four-point Likert-type scale ranging from 1 “not at all” to 4 “very much so” (scores range from 0-80). Gender-specific reference means are 36.5 (SD = 21.4) for men and 43.7 (12.6) for women, with cut-offs being 66 for men and 69 for women (Pasquali et al., 1994)

Volunteers of the present research answered the questionnaires in the Google Forms online platform that was configured in the same order of presentation: 1) Term of Consent, 2) demographic and attitudinal questionnaire, 3) PSS-10, 4) FDI, 5) S-STAI, 6) Thank you page. Those participants who answered “no” to the Term of Consent were addressed to the Thank you page without having any exposure to the other questionnaires. The first round of data collection (time 1) took place between March 20^th^ and March 25^th^, 2020, whereas the second round (time 2) happened between April 15^th^ and April 20^th^, 2020.

After data collection, Google Spreadsheets were utilized to consolidate the database and to export it in the .csv format. Then, researchers used SPSS (IBM, version 21.0) to run the analyses. Descriptive statistics of PSS-10, FDI and S-STAI were calculated for each categorical (demographic) variable with exception of those that were answered with the Likert-type scales. Due to the large amount of variables collected on an online platform, Cronbach’s alpha (α) was calculated for the three scales at time 1 and time 2; results were expected to show α > .70. To ensure the quality of data, beyond reliability, one-sample normality was tested using Kolmogorov-Smirnov analysis with the expectation non-significant results. Pairwise t-test comparisons between groups were computed to identify significant differences between the first round (time 1) and second round (time 2) of data collection for the whole sample. A repeated-measures ANOVA was performed to compare within and between groups for each demographic independent variable. Furthermore, prevalence of stress, depression and anxiety-like symptoms were calculated in percentage of participants above the means and cut-off points respective to the norms developed in previous studies in the Brazilian sample (Cacciari et al., 2016; Filgueiras et al., 2014; Pasquali et al., 1994)

A correlation matrix of the PSS-10, FDI and S-STAI results at time 1 and time 2 were calculated using Pearson correlation to identify possible discrepancies, significant associations, and validity of these measures. Additionally, exercise frequency at time 2 was included in the correlation matrix to identify significant associations with mental health variables. To assess correlations between continuous mental health scores and ordinal nutrition and exercise variables recorded at Time 2 (diet change for the better, weight gain, weight loss, and change in exercise frequency), Kendall rank correlations coefficients were calculated. For all correlations, significance was deemed when *p* < .05. The authors opted to compute three Linear Multiple Regressions (LMR) using the stepwise method to find the strength and ability of independent variables (i.e., demographic, behavioural and attitudinal) to predict PSS-10, FDI and S-STAI total scores at time 2. Total scores of mental health questionnaires at time 1 were put in the first step of the LMR, and the other variables were put in the second step. Categorical items were identified as dummy variables, whereas Likert-type answers were computed as ordinal data. The criterion for keeping a variable in the regression was the same as with other null-hypothesis tests (i.e., pairwise t-test and repeated-measures ANOVA); significance was deemed when *p* < 0.05. The coefficient beta (*β*) was inspected to reveal the direction and strength of the association between independent and dependent variables; whereas the coefficient of determination (*r*^*2*^) revealed the amount of variance explained by the model.

Finally, effect-sizes for the t-test of the LMR and the repeated-measures ANOVA (between, within and interaction) were calculated using the software G*Power 3.1, which also provided the interpretation criteria. The t-test effect-size was measured with Cohen’s d; the rule of thumb for this measure is: above 0.20 and below 0.50, the effect is small, above 0.50 and below 0.80, the effect is moderate, above 0.80 the effect is large. The repeated-measure ANOVA effect-size was measured by Cohen’s f and categorization goes as follows: above 0.10 and below 0.30, the effect is small, above 0.30 and below 0.50 the effect is moderate, and above 0.50 the effect is large.

## Results

Participants reported an age average of 37.90 (SD=12.33) years and were in quarantine for 3.52 (SD=1.77) days in the first round of data collection and 19.08 (SD=3.86) days in the second round. In terms of education, 98 volunteers reported to have either begun or finished high school (27.2%), 175 had either begun or finished began College (48.6%), 57 had either begun or finished a Master’s course (15.8%) and 30 (8.3%) had either begun or finished their PhD.

Participants reported changes in diet during the second round of data collection in reference to the first round. One hundred and sixteen participants (32.2%) reported to have worsened their diets from time 1 to time 2, 59 (16.4%) reported no significant changes in diet habits, whereas 185 (51.4%) answered that they were having a better diet than when they began quarantine.

At time 1, those reporting no exercise were 219 (60.8%), 1-to-3 days a week N=72 (20.0%) and 4 or more days a week N=69 (19.2%). At time 2, no exercise was 69 (19.2%), 1-to-3 days a week N=14 (3.9%) and 4 or more days of exercise N=277 (76.9%). No one reported exercising 7 days a week. This was in contrast to perceptions of change in exercise. One hundred eleven (30.8%) of respondents reported exercising less, 147 (40.8%) reported the same level of exercise and 102 (28.3%) reported more exercise. The percentage of women and men who did tele-psychotherapy was 72.8% and 27.2%, respectively.

Even though data collection used an online platform and participants had to answer a large amount of questions, Scales were reliable according to the adopted criterion (α > .70) in both time 1 and 2. The PSS-10 had α = .855 in the first round and α = .834 in the second round. The FDI presented α = .911 and α = .954 in times 1 and 2, respectively. The SSTAI showed α = .759 in the first time period and α = .713 in the second time period. Normality of continuous variables were also ensured in both Times 1 and 2. The PSS-10 presented a normal distribution in the first data collection with KS=1.189; *p* = .119, whereas in the second data collection it was KS=.836; *p* = .487. The FDI had the following results: Time 1 was KS=1.309; *p* = .065 and Time 2 was KS=1.093; *p* = .135. Finally, SSTAI had normality in Time 1 for KS=.978; *p* = .294 and Time 2 for KS=1.115; *p* = .166.

Table 1 depicts average and standard deviation (SD) of PSS-10, FDI and SSTAI stratified by the independent variables at time 1 and time 2. Mental health variables worsened significantly from the first round of data collection to the second, i.e., stress (*p* = .007), depression (*p* = .00003) and anxiety (p = .004). Repeated-measures ANOVA revealed that within-group effects were significant for all outcomes (criterion p < .05) demonstrating that from time 1 to time 2 there was an increase in stress, depression, and anxiety. Across all mental health outcome variables, significant between-group effects were observed for the following 3 predictors: 1. gender (women had significantly higher scores than men), 2. changes in diet (participants who felt that their diet worsened reported increased levels of psychological issues), and 3. amount of information (those who reported to receive too much information about COVID-19/quarantine also showed greater mental health dysfunction). Effect sizes ranged from .01 to .51. A gender x time interaction was observed (*p* < .001) for perceived stress. Men did not change in stress level but women had a significant increase (effect size = .28).

**Table 1.** Psychosocial variables: Perceive Stress, Depression and Anxiety symptoms by demographic and behavioural independent variables.

| Independent Variables | Psychosocial Variable |  |  |  |  |  |
| --- | --- | --- | --- | --- | --- | --- |
|  | Perceived Stress (PSS-10) |  | Depression (FDI) |  | State Anxiety (S-STAI) |  |
|  | Time 1 | Time 2 | Time 1 | Time 2 | Time 1 | Time 2 |
| *Whole Sample (N=360)* | 20.54 (6.99) | 22.03 (6.45)** | 65.32 (24.96) | 70.31 (25.44)*** | 43.61 (21.51) | 49.88 (20.13)** |
| *Gender* |  |  |  |  |  |  |
| Men (N=112) | 17.87 (7.59) | 17.28 (5.85) † | 54.98 (23.75) | 59.79 (25.42)** | 36.53 (21.52) | 45.87 (22.01)*** |
| Women (N=248) | 21.75 (6.35) | 24.18 (5.50)*** | 69.98 (24.12) | 75.06 (24.03)** | 46.81 (20.77) | 51.69 (18.98)** |
| *Education* |  |  |  |  |  |  |
| High School (N=98) | 20.94 (7.04) | 22.73 (6.19) * | 74.46 (23.67) | 80.91 (21.38)** | 46.40 (22.32) | 53.36 (20.56)*** |
| Bachelor Degree (N=175) | 22.33 (6.91) | 21.30 (6.38) * | 61.51 (24.67) | 67.27 (24.78)** | 41.55 (20.69) | 48.21 (19.46)*** |
| Master's Degree (N=57) | 21.39 (7.05) | 23.14 (6.39) † | 64.16 (24.22) | 65.05 (27.52) † | 45.82 (21.74) | 49.32 (20.24) † |
| Doctorate (N=30) | 19.74 (6.91) | 21.87 (7.49) † | 59.83 (25.53) | 63.43 (28.24) † | 42.30 (22.77) | 49.30 (21.83) * |
| *Changes in Diet* |  |  |  |  |  |  |
| For worse (N=116) | 21.23 (7.26) | 23.26 (6.61)*** | 72.78 (26.04) | 76.40 (23.52)** | 54.41 (22.24) | 67.47 (13.73)*** |
| No changes (N=59) | 21.05 (6.45) | 22.95 (5.53) * | 67.39 (24.15) | 71.12 (25.97) † | 46.76 (21.15) | 47.49 (17.15) † |
| For better (N=185) | 19.95 (6.96) | 20.97 (6.47) * | 59.98 (23.31) | 66.24 (25.77)*** | 35.83 (17.76) | 39.60 (16.63)** |
| *Changes in Exercise Frequency* |  |  |  |  |  |  |
| Fewer days (N=111) | 20.69 (7.79) | 21.29 (7.35) † | 63.19 (25.05) | 69.33 (25.65)** | 43.27 (22.96) | 51.41 (22.13)** |
| Same frequency (N=147) | 20.47 (6.60) | 21.97 (6.31)* | 65.71 (25.29) | 70.88 (25.32) * | 42.97 (21.00) | 48.67 (19.13) * |
| More days (N=102) | 20.48 (6.67) | 22.78 (5.52)** | 67.06 (24.96) | 70.55 (25.61) * | 44.90 (20.76) | 49.95 (19.31) * |
| *Days of Exercise* |  |  |  |  |  |  |
| 0 days (N=69) | 22.91 (7.70) | 26.41 (7.57)*** | 69.03 (25.24) | 74.58 (24.82)** | 48.23 (21.90) | 54.67 (20.52)** |
| 3 days (N=34) | 16.86 (7.47) | 17.40 (7.18) † | 62.71 (26.21) | 69.79 (26.08) * | 46.93 (25.72) | 55.43 (21.02) † |
| 4 days (N=135) | 21.38 (7.47) | 21.73 (5.75) † | 64.54 (25.07) | 69.25 (25.35) * | 43.26 (21.60) | 50.85 (19.58)*** |
| 5 days (N=80) | 19.88 (6.62) | 21.44 (4.83) * | 66.70 (23.18) | 70.60 (24.91) * | 41.92 (19.64) | 46.24 (18.48) * |
| 6 days (N=42) | 20.29 (5.34) | 22.21 (4.35) † | 59.29 (27.59) | 66.19 (25.44) * | 40.05 (23.06) | 45.67 (23.02)** |
| *Type of Exercise* |  |  |  |  |  |  |
| Aerobic (N=107) | 20.51 (7.01) | 20.74 (6.04) † | 59.36 (23.56) | 60.23 (26.28) † | 42.08 (21.07) | 47.21 (20.39) * |
| Anaerobic (N=122) | 20.00 (6.36) | 21.11 (5.46) * | 67.17 (24.85) | 75.21 (23.30)*** | 43.21 (21.25) | 50.04 (18.82)*** |
| Both (N=62) | 19.02 (6.80) | 21.21 (6.80) * | 67.81 (25.82) | 74.58 (24.82) * | 41.87 (22.14) | 48.81 (21.21) * |
| None (N=69) | 22.91 (7.70) | 26.41 (7.57)*** | 69.03 (25.24) | 74.58 (24.82)** | 48.23 (21.90) | 54.67 (20.52)** |
| *Amount of information* |  |  |  |  |  |  |
| Few information (N=18) | 20.78 (7.89) | 21.39 (6.56) † | 60.78 (28.74) | 66.17 (28.86) † | 34.11 (18.05) | 37.67 (19.05)** |
| Enough information (N=153) | 19.50 (6.76) | 20.75 (6.30) * | 61.52 (23.62) | 66.22 (26.54) * | 33.68 (17.63) | 37.26 (16.16)** |
| Too much information (N=189) | 21.36 (7.01) | 23.13 (6.39)** | 68.83 (25.26) | 74.02 (23.71)** | 52.55 (20.83) | 61.25 (16.10)*** |
| *Use of telemedicine* |  |  |  |  |  |  |
| Yes (N=40) | 22.05 (6.73) | 22.43 (5.16) † | 63.58 (24.82) | 66.88 (24.72) † | 41.05 (17.36) | 47.65 (17.89) * |
| No (N=320) | 20.35 (7.01) | 21.98 (6.60)** | 65.53 (25.01) | 70.74 (25.53)** | 43.93 (21.98) | 50.15 (20.40)*** |
| *Use of online nutrition* |  |  |  |  |  |  |
| Yes (N=86) | 20.89 (6.78) | 22.09 (6.42)** | 67.56 (24.83) | 72.11 (25.63)** | 44.89 (21.79) | 48.16 (19.77)** |
| No (N=274) | 19.43 (7.54) | 21.83 (6.58)** | 58.17 (24.15) | 64.59 (24.10)** | 39.52 (20.20) | 55.35 (20.39)*** |
| *Use of online fitness coaching* |  |  |  |  |  |  |
| Yes (N=142) | 20.97 (6.86) | 22.74 (6.59)** | 65.82 (24.85) | 70.82 (25.78)** | 43.17 (21.46) | 49.99 (20.27)** |
| No (N=218) | 19.89 (7.15) | 20.94 (6.09)** | 64.55 (25.20) | 69.53 (24.99)** | 44.28 (21.66) | 49.70 (19.97) * |
| *Use of telepsychotherapy* |  |  |  |  |  |  |
| Yes (N=136) | 20.57 (6.52) | 21.94 (6.24)** | 61.88 (23.12) | 64.72 (24.04) * | 42.54 (21.12) | 47.46 (19.67)** |
| No (N=224) | 20.52 (7.27) | 22.08 (6.59)** | 70.97 (26.87) | 79.51 (25.09)** | 45.38 (22.11) | 53.85 (20.30)** |
| *Risk for COVID-19* |  |  |  |  |  |  |
| Yes (N=98) | 23.59 (6.79) | 27.14 (6.14)** | 66.49 (25.21) | 72.32 (25.58)** | 48.96 (21.10) | 58.74 (18.36)** |
| No (N=262) | 19.40 (6.73) | 20.12 (5.45) * | 64.88 (24.90) | 69.56 (25.40)** | 41.61 (21.36) | 46.56 (19.78)** |

Note: \* $p < 0.05$ ; \*\* $p < 0.01$ ; \*\*\* $p < 0.001$ ; † non-significant difference

Beyond those three variables, between groups significant differences regarding perceived stress occurred for four other variables: number of days of exercise per week, type of exercise, use of online fitness coaching and risk for COVID-19. Regarding symptoms of depression, statistical differences appeared for four other variables: education, type of exercise, use of online nutritionists and use of tele-psychotherapy. Finally, regarding state anxiety, between groups significant differences were shown for two other variables: risk for COVID-19 and use of tele-psychotherapy. Results for the repeated-measures ANOVA are depicted in the supplemental material.

For perceived stress, 237 (65.8%) and 269 (74.7%) of participants scored above the population mean at time 1 and 2, respectively. Prevalence of excessive stress (≥2 SD above reference mean) was 6.9% (IC 95 5.2%-8.6%) in the first round and 9.7% (IC 95 8.2%-11.2%) in the second round. Of the 34 individuals in this category, 94% of these individuals were women. 82% did no exercise at all, but the remaining 18% reported 6 days a week of exercise. Also, 0% utilized tele-psychotherapy. Regarding symptoms of depression, 224 (62.2%) and 260 (72.2%) of participants were above the reference mean at Time 1 and 2, respectively. High levels of symptoms of depression (≥2 SD above reference mean) had a prevalence of 4.2% (IC 95 3.6%-4.8%) at time 1 and 8.0% (IC 95 7.1%-8.9%) at time 2. Participants > 2 SD (n = 24) were mostly women (88%) and did not utilize tele psychotherapy (88%). The number of male participants above the reference mean for state anxiety was 54 (48.2%) and 72 (64.3%) at time 1 and 2, respectively. For women it was 132 (53.2%) and 163 (65.7%). Prevalence of excessive state anxiety (≥2 SD above reference mean) was 8.7% (IC 95 7.4%-10.0%) in the first round against 14.9% (IC 95 12.3%-17.5%) in the second round. Those > 2 SD had worsening diet (45 of 53 participants) and reported no tele-psychotherapy (81%).

All mental health outcome variables had significant positive correlations with each other, which can be seen in Figure 1. The strongest correlations were observed in intertemporal correlations of the same variable (*r* = .61 for PSS-10, *r* = .79 for FDI, and *r* = .69 for STAI-S). Small to moderate significant associations were seen between different mental health variables (r = .13 - .50). Higher exercise frequency at Time 2 had negative correlations with all mental health variables, which was significant for stress and anxiety, but not for depression. Kendall rank correlations were conducted to test the association between continuous mental health variables, and ordinal nutritional and exercise variables collected at Time 2 (diet change, weight gain, weight loss, and change in exercise frequency). Diet change was coded so that a higher score reflects a diet change for the better. It was found that positive change in diet had a negative association with FDI Time 1 (*τ* = -.18, *p* = .01), FDI Time 2 (*τ* = -.14, *p* = .01), STAI Time 1 (*τ* = -.30, *p* = < .001), and STAI Time 2 (*τ* = -.49, *p* < .001). No significant associations were found between weight loss at Time 2 and perceived stress at any time. No significant associations were found between mental health outcome variables and weight loss or change in exercise frequency, at any time, with the exception of a negative association between greater weight loss and STAI Time 2 (*τ* = -.36, *p* = .026)

**Figure 1.**
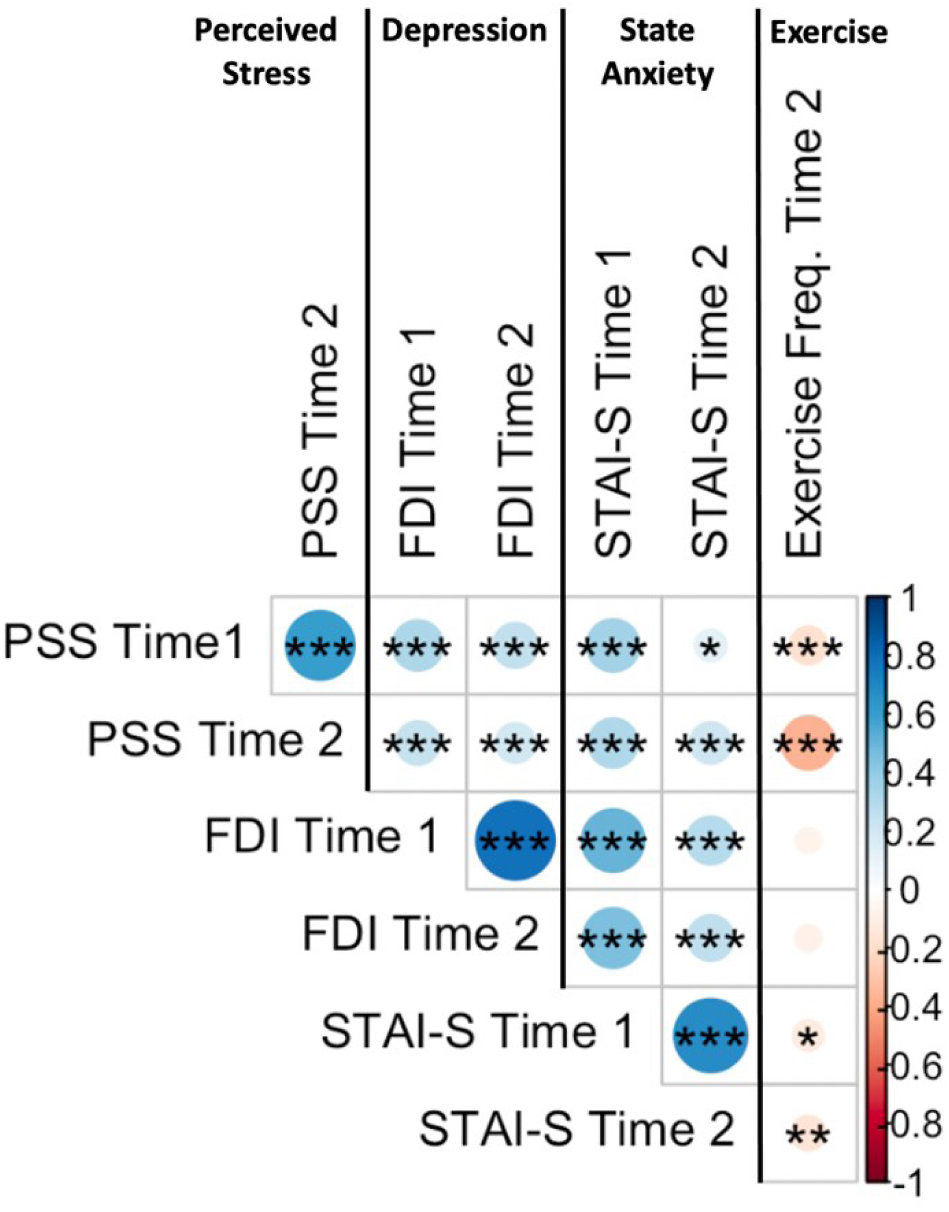
Pearson correlation of psychosocial variables (Perceive Stress, Depression and Anxiety) at Time 1 and Time 2, and exercise frequency at Time 2. Time 1 comprises data collection between March 20^th^ and 25^th^, while Time 2 entails data collection between April 15^th^ and 20^th^. Size and color of circle indicate the strength and direction of correlation. * < *p* .05, ** *p* < .01, *** *p* < .001

The linear multiple regression (LMR) model for perceived stress revealed that the dependent variable (PSS-10, time 2) was predicted by the score of the PSS-10 at time 1, number of days of exercise, risk for COVID-19, types of exercise, changes in the frequency of exercise, feeling imprisoned, days in quarantine and gender in order of strength of the coefficient β. Altogether, those variables explained 56% of the variance. The symptoms of depression LMR showed that the dependent variable (FDI time 2) was predicted by the score of the FDI time 1, types of exercise, own ability to understand what is happening, level of education and gender respectively. The independent variables explained 33% of the variance of depression in the second round of data collection. Finally, the state anxiety LMR depicted that the dependent variable (S-STAI time 2) was predicted, in order of association, risk for COVID-19, feeling safe, the score of S-STAI time 1, weight loss, changes on diet, amount of COVID-19 information, feeling imprisoned and age. The independent variables of this LMR explained cumulatively 42% of the variance. Table 2 presents the coefficient β, the t-test statistics, effect-size and coefficient of determination for the three LMR.

**Table 2.** Separate stepwise LMR using demographic and behavioural variables to predict Perceived Stress, Depression and Anxiety symptoms. The table provides the coefficient of regression (*β*), the t-test and effect-size. Additionally, results present the coefficient of determination (r^2^) to assess the amount of variance explained by models.

| Variable | Multiple Linear Regression Statistics |  |  |  |
| --- | --- | --- | --- | --- |
| | $\beta$ | t-test | effect-size | $r^2$ |
| Perceived Stress (PSS-10) time 2 |  |  |  | 0.56 |
| (Intercept) | 1.13 | 0.67 | 0.20 |  |
| PSS-10 time 1** | 3.78 | 4.98 | 0.62 |  |
| Gender* | 0.35 | 2.03 | 0.25 |  |
| Feeling Imprisoned* | 1.28 | 1.97 | 0.16 |  |
| Days in quarantine* | 0.75 | 2.02 | 0.20 |  |
| Days of exercise** | 3.46 | 3.38 | 0.53 |  |
| Types of exercise** | 2.41 | 3.95 | 0.56 |  |
| Risk for COVID-19** | 2.87 | 4.77 | 0.59 |  |
| Changes in freq. Exercise* | -1.54 | 2.01 | 0.13 |  |
| Depression (FDI) time 2 |  |  |  | 0.33 |
| (Intercept)** | -9.02 | 4.97 | 0.60 |  |
| FDI time 1** | 3.75 | 5.76 | 0.42 |  |
| Gender* | 0.48 | 1.98 | 0.18 |  |
| Types of exercise** | 2.54 | 3.41 | 0.34 |  |
| Understanding* | -1.87 | 1.99 | 0.19 |  |
| Education* | -0.96 | 2.08 | 0.33 |  |
| State Anxiety (S-STAI) time 2 |  |  |  | 0.42 |
| (Intercept) | -9.41 | 1.14 | 0.40 |  |
| S-STAI time 1** | 3.41 | 4.71 | 0.37 |  |
| Change on diet** | -2.65 | 3.62 | 0.14 |  |
| Weight Loss** | 2.90 | 2.01 | 0.10 |  |
| Age* | 0.20 | 3.59 | 0.50 |  |
| Feeling Safe** | -3.90 | 3.89 | 0.44 |  |
| Feeling Imprisoned* | 1.05 | 2.59 | 0.70 |  |
| Risk for COVID-19** | 4.75 | 3.65 | 0.34 |  |
| Amount of information* | 2.18 | 1.98 | 0.11 |  |

Note: \* $p < 0.05$ ; \*\* $p < 0.01$

## Discussion

The current investigation provides a unique glimpse into the mental health of Brazilians at the beginning of government-mandated quarantine from the COVID-19 pandemic, a novel, disruptive and society-wide stressor. Findings indicate that a substantial portion of respondents were distressed at both time points, with worsening mental health from the initiation of quarantine to a point a few weeks later. More specifically, increases in perceived stress, symptoms of depression and state anxiety were observed, with a gender by time interaction recorded for stress. Men experienced increases in depression and anxiety over time, but not for perceived stress. Across genders, the number of days in quarantine was linearly related to worse perceptions of perceived stress. Repeated measures ANOVA revealed that 3 factors were all related to worse levels of stress, depression and anxiety: female gender, worsening diet and excess of COVID-19 information. In regression analyses, however, mental health outcomes were associated with a variety of other demographic, COVID-19 specific, and behavioural factors, such as use of tele-psychotherapy. Exercise-related factors, such as exercise frequency, were the predominate predictors of perceived stress.

A substantial portion of the participants reported levels of stress, depression and anxiety above established means for the population. At time 2, greater than 70% of the sample was above the normative mean for both stress and depression. For anxiety, >60% of both men and women were above the normative mean. More importantly, some participants scored very high for mental health disturbances, especially at time 2. For stress, 9.7% of the sample was above 2 SD at time 2, whereas the prevalence according to the Brazilian norms is 6.8% (Cacciari et al., 2016). This was an increase from 6.9% at time 1. Similar trends were seen for depression (4.2% at time 1, 8.0% at time 2; versus a norm of 4.1%) (Filgueiras et al., 2014) and state anxiety (8.7% increasing to 14.9%; versus a norm of 9.4%) (Pasquali et al., 1994). This is similar to anxiety levels observed in a large sample during quarantine in China (Hu et al., 2020). While the percentage of individuals scoring at these extremes is still relatively low, it potentially represents a huge increase in burden to society when multiplied across the entire population. Mental health initiatives on the national level would have to be scaled up to meet new demand (Funk et al., 2008). Key to this endeavour would be: a) identifying those most at risk and b) properly assessing their condition.

In the effort to identify those most at risk, pertinent predictors of mental health outcomes were analysed. Interestingly, each mental health indicator was predicted by a varying set of factors. Anxiety was predicted by the greatest number of COVID-19 related factors: feelings of safety, feelings of being imprisoned, risk for COVID-19 and amount of information. In other words, those who felt unsafe, cooped up, at risk for infection and being inundated with information demonstrated higher levels of anxiety. This falls in line with an expansive literature reporting that feelings of anxiety burgeon when people feel under threat, unsafe, have too many options and an uncertain future (Carretta et al., 2014; Gilbert et al., 2008). Depression, a condition which can be usually seen as compromised of regrets from the past (Buechler, 2015), was understandably not predicted by COVID-19 related factors. Only “understanding what is happening” was a significant inverse predictor. Stress was predicted by feelings of being imprisoned, days in quarantine and risk for COVID-19 and also by a number of exercise factors.

Other similar longitudinal studies during the COVID-19 pandemic also found similar results. For example, O’Connor et al. (2020) gathered information about mental health and wellbeing among adults in UK during lockdown. They found that suicidal ideation increased over time. Additionally, symptoms of anxiety increased and those with pre-existing mental health problems showed worse mental health outcomes, longitudinally. On the other hand, symptoms of depression were stable; nonetheless, it is important to highlight that suicidal ideation is a symptom of depression and the scale adopted by O’Connor et al. (2020) is a 9-item brief self-report questionnaire based on DSM-IV, whereas the FDI adopted in the present research is a 30-item psychometric measure based on DSM-V. Differences in number of items, items structure and diagnosis criteria are potential explanations for the differences between O’Connor et al. (2020) findings and this study regarding depressive symptoms. A longitudinal cohort study from UK also suggests that general mental health deteriorated when compared to the period before the COVID-19 pandemic (Pierce et al., 2020); however, there were no specificities regarding different types of psychiatric symptoms. Their findings corroborate to this study regarding gender and age: younger women tend to show poorer mental health.

A study by Canet-Juric et al. (2020) some conflicts with the findings of the present study, even though their sample comes from Argentina, a Brazilian neighbour in South America. Therefore, the cultural and social conditions seem to play an important role in mental health. They found that depression increased through time, differently from the stability found by O’Connor et al. (2020), though anxiety decreased. In comparison to the results from Canet-Juric et al. (2020), the present study corroborates to an increase of the symptoms of depression, but it goes in the opposite direction regarding symptoms of anxiety. Longitudinal studies (Canet-Juric et al. (2020); O’Connor et al. (2020); Pierce et al. (2020)) suggest changes in levels of symptoms in mental health during the COVID-19 pandemic, however, there is no common ground so far and future studies should consider cultural and social variables to explain this phenomenon.

In general, exercise was associated with mental health outcomes in the expected manner – more frequent exercise and aerobic exercise were related to the lowest levels of distress. For all 3 mental health outcomes, those with no exercise (0 days per week) had the highest average levels of stress (22.9 at time 1 to 26.4 at time 2), depression (69.0 to 74.6) and anxiety (48.2 to 54.7). These seems to support the previous findings that “something is better than nothing” (Ekkekakis et al., 2000; Werneck et al., 2018). In linear regression, perceived stress was related to the greatest number of exercise-related factors: exercise frequency per week, type of exercise and perceived changes in exercise behavior. Higher frequency of exercise (days/week) was associated with less stress. However, the linear relationship between perceived stress and exercise frequency was small (r = -.28), which is line with previous investigations (Stults-Kolehmainen and Sinha, 2014). It should be noted that 58.2% of the sample reported that they perceived that their exercise behaviour changed within a few weeks of quarantine (30.8% doing less and 28.3% doing more), which follows the known phenomenon that stressful events can either inhibit or activate changes in exercise behaviors (Stults-Kolehmainen and Sinha, 2014). Furthermore, those who perceived that they exercised more frequently from Time 1 to Time 2 had less stress. Interestingly, of those very high for stress (< 2SD), 82% do no exercise at all, but the remaining 18% complete 6 days a week of exercise. In LMR analysis, exercise factors explained 13.1% of the adjusted variance in stress. It has been hypothesized that a lack of physical activity can lead to feelings of tension for those accustomed to it. Furthermore, stressful circumstances may activate desires, wants, or urges to move and be active in some individuals (Stults-Kolehmainen et al., 2020). For repeated measures, the results were slightly different, with changes in exercise not being significant, but use of online fitness coaching reaching significance. An interaction was observed in that those who performed aerobic exercise had the lowest levels of depression at both time points. In fact, those who did aerobic exercise did not have any increase in depression. However, the clearest association of exercise frequency and mental health was for anxiety. Those at the highest levels of exercise had the lowest anxiety and each day less was associated with more anxiety.

Aside from exercise, there were notable findings for dietary habits and use of tele-psychotherapy. Those who rated their dietary habits as becoming worse also had higher levels of stress, depression and anxiety. Those with the highest levels of anxiety were those with worsening diet at the second time point (effect size for interaction was .37). Those who used online nutrition services had lower levels of depression, but there was no difference for stress or anxiety. Those who utilized online psychotherapy reported lower levels of depression and anxiety. While there is no income data to explain use of online resources, those using online resources were more educated. Thus, one might surmise that those from better off demographic groups are less affected partly because of greater access to resources. Given the limited quantity of resources to mitigate mental health impairments during crises, such as pandemic and quarantine, it is crucial to identify the risk factors that may predispose individuals for worsening outcomes. Further, it is crucial to allocate or develop resources that can have a wide reach to affected populations.

### Limitations of the Study

Despite the progress this study makes in tracking changes in mental health and identifying risk factors, the current research does demonstrate some limitations. First, there was no pre-quarantine baseline and assessments spanned less than a single month. Furthermore, validated measures of exercise and dietary habits, which can be very lengthy, were not utilized to reduce survey fatigue. More importantly, the current data needs interpreted with some caution because factors other than quarantine could contribute to changes in the mental health outcomes observed, such as growing political and economic unrest in Brazil (Prado, 2020). At the time of data collection, Brazil was going through political and social changes with a far-right government and our results must be seen in perspective. The first reaction of the Brazilian president towards COVID-19 was denial and down-play of the seriousness of the virus. Further, public policies were adopted based on conspiracy theories (Barberia and Gómez, 2020). The political view of the Brazilian leader has influenced the population’s perspective towards quarantine. A study from Ramos et al. (2020) showed that Brazilians who share the same political beliefs with the president tend to trust in non-scientific assumptions that reduces their own perception of risk. This distrust leads to less careful behaviours and may have influenced the present results. The fact that this study did not consider political views constitute a limitation that should be considered. Also, it should be noted that effect sizes for changes within approximately three weeks were small (Cohen’s d were .25 – stress, .30 – depression, and .38 – anxiety), possibly because in some cases individuals had improved mental health (n = 31; 8.6%) due to quarantine conditions, such as being closer to loved ones throughout the day or being removed from dangerous work environments. This sample may be considered small compared to other longitudinal studies (e.g., Canet-Juric et al. (2020); O’Connor et al. (2020); Pierce et al. (2020)), and therefore, may exhibit less robust inferential analyses - in terms of reliability and normality of data. Lastly, the data was self-reported as it was collected online on Google Forms. Finally,

## Conclusion

This study found that mental health worsened following a government-mandated quarantine in Brazil. These data are unique because observations were made in the first days and weeks after quarantine was decreed due to the COVID-19 pandemic crisis. Specifically, from the time point when quarantine was decreed until 1 month later, worsening perceived stress, symptoms of depression and anxiety was observed in this sample of the Brazilian population. Analyses from this study identified several risk factors for mental health, including gender (being female), lower education, less frequent exercise, worsening diet and a lack of resources, such as access to tele-psychotherapy. COVID-19 related factors predicted anxiety and stress more so than symptoms of depression. The implications of these data are clear; mental health worsens, requiring more resources to improve the experience of life in quarantine. The extent to which these can be diligently developed and allocated will depend on a data-driven process that begins with monitoring and analysing data as we and others have initiated.

## Data Availability

Ethical Committee did not allow data availability to preserve possible participant identification.

## Authorship

A.F. designed the study and collected data. M.B. and A.F. performed analysis. A.F. and M.S.K. drafted the manuscript. M.B assisted with interpretation of result and provided critical revisions. All authors approved of the final version of the paper for submission.

## Acknowledgements

The present research is funded by the Coordenacao de Aperfeicoamento de Pessoal de Nivel Superior (CAPES) of the Ministry of Education of Brazil under the PROAP program.

## Supplemental Material 1

Results of repeated-measures ANOVA with F-statistics, degree of freedom (df), p-value and 636 effect size for each psychosocial factor (stress, depression and anxiety) by demographic and 637 behavioural variables separately.

| Variable | Repeated-measures statistics |  |  |  |  |  |  |  |  |  |  |  |
| --- | --- | --- | --- | --- | --- | --- | --- | --- | --- | --- | --- | --- |
|  | Perceived Stress (PSS-10) |  |  |  | Depression (FDI) |  |  |  | State Anxiety (S-STAI) |  |  |  |
|  | *F*-statistics | df | *p*-value | effect-size | *F*-statistics | df | *p*-value | effect-size | *F*-statistics | df | *p*-value | effect-size |
| *Gender* |  |  |  |  |  |  |  |  |  |  |  |  |
| Between | 74.50 |  | <0.001 | 0.28 | 33.97 |  | <0.001 | 0.09 | 14.19 |  | <0.001 | 0.11 |
| Within | 7.70 | 1.358 | <0.010 | 0.23 | 28.17 | 1.358 | <0.001 | 0.29 | 57.61 | 1.358 | <0.001 | 0.35 |
| Interaction | 20.74 |  | <0.001 | 0.53 | 0.02 |  | =0.882 | 0.01 | 5.67 |  | <0.050 | 0.03 |
| *Education* |  |  |  |  |  |  |  |  |  |  |  |  |
| Between | 1.91 |  | =0.127 | 0.02 | 8.29 |  | <0.001 | 0.06 | 1.50 |  | =0.214 | 0.03 |
| Within | 9.08 | 1.356 | <0.005 | 0.17 | 15.73 | 1.356 | <0.001 | 0.13 | 31.63 | 1.356 | <0.001 | 0.20 |
| Interaction | 1.21 |  | =0.307 | 0.01 | 1.66 |  | =0.175 | 0.02 | 0.64 |  | =0.589 | 0.01 |
| *Changes on Diet* |  |  |  |  |  |  |  |  |  |  |  |  |
| Between | 3.70 |  | <0.050 | 0.13 | 8.75 |  | <0.001 | 0.18 | 74.06 |  | <0.001 | 0.37 |
| Within | 22.13 | 1.357 | <0.001 | 0.29 | 22.42 | 1.357 | <0.001 | 0.12 | 39.45 | 1.357 | <0.001 | 0.51 |
| Interaction | 1.17 |  | =0.311 | 0.04 | 1.14 |  | =0.320 | 0.04 | 16.50 |  | <0.001 | 0.25 |
| *Changes on Frequency of Exercise* |  |  |  |  |  |  |  |  |  |  |  |  |
| Between | 0.54 |  | =0.581 | 0.02 | 0.35 |  | =0.703 | 0.01 | 0.29 |  | =0.748 | 0.01 |
| Within | 21.66 | 1.357 | <0.001 | 0.13 | 32.01 | 1.357 | <0.001 | 0.11 | 50.78 | 1.357 | <0.001 | 0.17 |
| Interaction | 1.21 |  | =0.298 | 0.04 | 0.72 |  | =0.490 | 0.03 | 1.07 |  | =0.345 | 0.03 |
| *Number of days of exercise per week* |  |  |  |  |  |  |  |  |  |  |  |  |
| Between | 12.17 |  | <0.001 | 0.32 | 1.06 |  | =0.378 | 0.04 | 2.19 |  | =0.070 | 0.08 |
| Within | 13.99 | 1.355 | <0.001 | 0.37 | 23.16 | 1.355 | <0.001 | 0.16 | 30.11 | 1.355 | <0.001 | 0.23 |
| Interaction | 3.21 |  | <0.050 | 0.09 | 0.34 |  | =0.851 | 0.01 | 0.64 |  | =0.634 | 0.02 |
| *Type of exercise* |  |  |  |  |  |  |  |  |  |  |  |  |
| Between | 9.68 |  | <0.001 | 0.21 | 6.01 |  | <0.001 | 0.22 | 1.95 |  | =0.121 | 0.08 |
| Within | 29.53 | 1.356 | <0.001 | 0.18 | 31.73 | 1.356 | <0.001 | 0.11 | 48.16 | 1.356 | <0.001 | 0.19 |
| Interaction | 4.78 |  | <0.005 | 0.11 | 3.82 |  | <0.050 | 0.08 | 0.25 |  | =0.862 | 0.01 |
| *Amount of information* |  |  |  |  |  |  |  |  |  |  |  |  |
| Between | 81.34 |  | <0.001 | 0.11 | 5.39 |  | <0.050 | 0.09 | 4.65 |  | <0.050 | 0.09 |
| Within | 13.82 | 1.357 | <0.001 | 0.13 | 5.45 | 1.357 | <0.050 | 0.10 | 12.92 | 1.357 | <0.001 | 0.48 |
| Interaction | 4.37 |  | <0.050 | 0.12 | 0.54 |  | =0.585 | 0.06 | 0.042 |  | =0.958 | 0.01 |
| *Use of telemedicine* |  |  |  |  |  |  |  |  |  |  |  |  |
| Between | 1.12 |  | =0.290 | 0.04 | 0.53 |  | =0.467 | 0.03 | 0.70 |  | =0.402 | 0.03 |
| Within | 4.00 | 1.358 | <0.050 | 0.09 | 9.63 | 1.358 | <0.001 | 0.11 | 21.26 | 1.358 | <0.001 | 0.19 |
| Interaction | 1.56 |  | =0.212 | 0.03 | 0.48 |  | =0.487 | 0.03 | 0.02 |  | =0.893 | 0.01 |
| *Use of online nutrition* |  |  |  |  |  |  |  |  |  |  |  |  |
| Between | 1.35 |  | =0.246 | 0.06 | 8.39 |  | <0.050 | 0.09 | 0.15 |  | =0.700 | 0.08 |
| Within | 23.86 | 1.358 | <0.001 | 0.18 | 29.48 | 1.358 | <0.001 | 0.25 | 96.91 | 1.358 | <0.001 | 0.28 |
| Interaction | 2.61 |  | =0.107 | 0.06 | 0.86 |  | =0.355 | 0.03 | 41.93 |  | <0.001 | 0.35 |
| *Use of online fitness coaching* |  |  |  |  |  |  |  |  |  |  |  |  |
| Between | 4.95 |  | <0.050 | 0.08 | 0.25 |  | =0.619 | 0.03 | 0.04 |  | =0.841 | 0.02 |
| Within | 19.25 | 1.358 | <0.001 | 0.14 | 32.02 | 1.358 | <0.001 | 0.21 | 46.91 | 1.358 | <0.001 | 0.32 |
| Interaction | 1.23 |  | =0.268 | 0.04 | 0.01 |  | =0.988 | 0.01 | 0.61 |  | =0.436 | 0.03 |
| *Use of telepsychotherapy* |  |  |  |  |  |  |  |  |  |  |  |  |
| Between | 0.01 |  | =0.944 | 0.01 | 22.49 |  | <0.001 | 0.22 | 4.99 |  | <0.050 | 0.07 |
| Within | 20.29 | 1.358 | <0.001 | 0.26 | 42.18 | 1.358 | <0.001 | 0.31 | 55.87 | 1.358 | <0.001 | 0.40 |
| Interaction | 0.09 |  | =0.765 | 0.06 | 10.59 |  | <0.001 | 0.33 | 3.93 |  | <0.050 | 0.07 |
| *Risk for COVID-19* |  |  |  |  |  |  |  |  |  |  |  |  |
| Between | 74.48 |  | <0.001 | 0.37 | 0.60 |  | =0.440 | 0.02 | 19.60 |  | <0.001 | 0.10 |
| Within | 37.98 | 1.358 | <0.001 | 0.18 | 29.46 | 1.358 | <0.001 | 0.13 | 57.28 | 1.358 | <0.001 | 0.47 |
| Interaction | 16.74 |  | <0.001 | 0.19 | 0.35 |  | =0.555 | 0.03 | 6.17 |  | <0.050 | 0.09 |

Note: Cohen's *f* interpretation is as follows above 0.10 and below 0.25, small effect-size, above 0.25 and below 0.40, moderate effect size, and above 0.40, large effect-size.

